# Quantitative interferon gamma responses to *Mycobacterium tuberculosis* in a community-based survey of adolescents and adults in Blantyre, Malawi

**DOI:** 10.64898/2026.04.02.26349931

**Authors:** Mphatso D. Phiri, Hannah M. Rickman, Christine Mandalasi, Aaron Chirambo, Kondwani C. Jambo, Henry C. Mwandumba, Elizabeth L. Corbett, Katherine C. Horton, Marc Y.R. Henrion, Peter MacPherson

## Abstract

**Background:** Binary interpretation of *Mycobacterium tuberculosis* (Mtb) interferon gamma release assay (IGRA) results discards information about recency of exposure and disease risk. We analysed quantitative IGRA responses to Mtb in a community-based survey to investigate associations with response magnitude and inform understanding of transmission dynamics.

**Methods:** We included QuantiFERON-TB Gold Plus (QFT-Plus) results from 2,895 participants (10-40 years old) in Blantyre, Malawi. Bayesian regression models assessed the probability of a positive response (≥0.35 IU/mL), response magnitude, and associated factors. We also investigated associations with a TB2-TB1 differential >0.6 IU/mL (proposed to reflect recent transmission), and how hypothetical alternative IGRA positivity thresholds affected inference about age- and sex-specific transmission.

**Results:** 17.4% (503/2,895) of participants had positive TB1 or TB2 responses at the QFT-Plus positivity threshold (0.35 IU/mL). The distributions of TB1 and TB2 responses, among participants with positive QFT-Plus, were similar across age and sex. A TB2-TB1 differential >0.6 IU/mL occurred in 3.8% (109/2,895) of participants and was not associated with age or sex. However, participants with HIV had reduced odds of TB2-TB1>0.6 IU/mL (adjusted odds ratio 0.37 [0.14-0.93]). At higher hypothetical positivity thresholds, the mean predicted Mtb immunoreactivity prevalence among males exceeded that in females at an earlier age: at 19 years, predicted immunoreactivity prevalence ratios were 0.90 (0.83-0.99) and 1.02 (0.89-1.15) at 0.1 IU/mL and 0.5 IU/mL thresholds, respectively.

**Conclusions:** Quantitative IGRA responses offer information about age- and sex-specific immunoreactivity and transmission risks that dichotomisation obscures. In high-burden settings, quantitative IGRA responses may clarify Mtb transmission patterns and guide targeted public health strategies.

**Main point:** We found high prevalence of *Mycobacterium tuberculosis* immunoreactivity (QuantiFERON-TB Gold Plus [QFT-Plus] positivity), in a community random sample of 10–40-year-olds in Blantyre, Malawi. Quantitative QFT-Plus responses showed an association between TB2-TB1>0.6 IU/mL (possible recent Mtb exposure) and HIV positivity.

## Introduction

Tuberculosis (TB) remains the world’s leading killer from a single infectious disease.[1] People exposed to *Mycobacterium tuberculosis* (Mtb) may progress to disease months, years, or even decades after exposure,[2,3], with the risk of progression decreasing with time since exposure.[3] In high transmission settings, incidence is driven by recent infections.[4] Therefore, identifying and protecting those recently infected at the highest risk of progression is key to reducing incidence and interrupting transmission to achieve global targets.[5]

Two main tests for identifying individuals exposed to Mtb are skin-based tests and blood-based interferon-gamma release assays (IGRAs). These tests elicit immunological memory responses in previously sensitised individuals.[6] Broadly, IGRAs have superior specificity for Mtb compared to the standard tuberculin skin test (TST), particularly in Bacille Calmette Guerin (BCG)-vaccinated populations and settings with high prevalence of non-tuberculous mycobacteria,[7] although newer skin tests perform similarly.[8] IGRAs have therefore become central to epidemiological research and vaccine trials in high-burden settings.[9] Yet both skin-based and IGRA tests share two fundamental limitations: they cannot resolve recent from remote infection, and poorly predict incident TB disease.[7]

Current IGRA tests are optimised for identifying individuals exposed to Mtb and at risk of disease, to guide preventive therapy. Most IGRAs dichotomise continuous responses, classifying individuals as positive or negative, at a fixed threshold, for example, 0.35 IU/mL for QuantiFERON assays.[7] The positivity threshold represents a trade-off between sensitivity and specificity estimated against TB disease as a reference.[10] However, the usefulness of IGRAs beyond individual disease risk stratification and current binary thresholds remains unknown. Recent evidence suggests that the magnitude of quantitative IGRA responses contains additional information about exposure and disease risk beyond binary classification.[11,12] In contact studies, higher interferon-gamma concentrations correlate with markers of intense or prolonged exposure, for example, contact time and sleeping in the same room with a person diagnosed with TB.[13] A systematic review of longitudinal cohorts found a dose-response relationship between baseline IGRA values and subsequent TB risk, rising steeply between 0.35 and 5 IU/mL, then plateauing around 15 IU/mL.[12] The relative risk of incident TB at 5 IU/mL was eleven-fold higher than at the 0.35 IU/mL threshold.[12] Moreover, it is plausible that responses below the manufacturer’s threshold represent true Mtb sensitisation without disease, or with disease not detected by current tests. The newest QuantiFERON assay, QFT-Plus, adds a second antigen tube designed to stimulate CD8+ T cell responses, which predominate in recent Mtb infection and incident disease.[11,13] A recent systematic review concluded that a greater TB2 response compared to TB1 correlates with infection risk and disease state.[14] Some investigators have proposed that high TB2 responses are associated with incident disease[15], whereas others propose that a TB2-TB1 differential exceeding 0.6 IU/mL indicates recent exposure,[13] not simply assay variability.[16] The evidence, however, is inconsistent: studies in low-incidence settings report associations with exposure intensity,[13] while those in high-burden populations find little discrimination between recent and remote exposure.[17]

Most studies that have analysed quantitative IGRA responses included participants purposively selected for their Mtb exposure risk (e.g. contacts of persons with TB, or individuals with low exposure risk, typically in low-burden countries). How quantitative IGRA responses are distributed in a general population with heterogeneous exposure in a high-burden setting, and which individual characteristics shape these responses, is unknown. We therefore analysed community-based IGRA survey data[18] to describe the distribution of interferon-gamma concentrations among adolescents and adults. We investigated whether responses increased with age, reflecting cumulative exposure, and whether young men showed higher values, given their elevated TB exposure risk.[19,20] We also investigated associations with a TB2-TB1 differential exceeding >0.6 IU/mL, and assessed how hypothetical alternative thresholds would alter Mtb immunoreactivity prevalence estimates and, by extension, inference about transmission intensity.

## Materials and methods

### Study setting and population

This is a secondary analysis of data from a community-based survey conducted in Blantyre City, Malawi.[18] Blantyre City, with an estimated population of ∼900,000 in 2024, has a high TB burden: adult prevalence of bacteriologically confirmed pulmonary TB was 150/100,000 in 2019.[21]

The survey aimed to investigate sex differences in the prevalence of Mtb immunoreactivity among adolescents and adults aged 10-40 years old. Participants were recruited from randomly selected households in 33 high TB burden neighbourhoods between 17^th^ January 2023 and 24^th^ March 2024. All household members aged 10-40 years who were present at the time of the study team’s visit were eligible and invited to participate. No additional exclusion criteria were defined. Mtb immunoreactivity was measured using the QuantiFERON- TB Gold Plus (QFT-Plus) assay. Detailed methods have been published previously.[18,22]

### Laboratory procedures

The QuantiFERON-TB Gold Plus (QFT-Plus – QIAGEN, Hilden, Germany) assay measures interferon gamma (IFN-γ) released by T cells following stimulation with Mtb peptides in two antigen tubes, TB1 and TB2. TB1 measures CD4^+^ T cell responses, with TB2 measuring both CD4^+^ and CD8^+^ responses.

Two additional tubes serve as negative and positive controls. The nil tube measures background, i.e. other infectious or sterile IFN-γ release, and the mitogen tube elicits non-specific IFN-γ release using phytohemagglutinin, acting as a positive control.

Blood collection and processing followed manufacturer instructions.[10] 5mL venous blood samples were drawn into 6 mL lithium heparin tubes, transported to a TB research laboratory at ambient temperature within 8 hours of collection. Blood was then pipetted into QFT-Plus nil, TB1, TB2 and mitogen tubes, and incubated at 37°C for 20 hours. After incubation, samples were centrifuged, and plasma was harvested and stored at -20^°^C.

IFN-γ concentrations were quantified within two weeks using a manual enzyme-linked immunosorbent assay (ELISA). Optical density values from the ELISA were converted to IFN-γ concentrations using a reference standard curve generated by the QFT-Plus software (v2.7.1.2). Quantitative IFN-γ values for the nil, TB1, TB2, and mitogen tubes were extracted, alongside QFT-Plus test results (positive, negative, or indeterminate). Participants with indeterminate results were included in the analysis, as the aim was to describe TB1 and TB2 responses, irrespective of the QFT-Plus result. IFN-γ values exceeding 10 IU/mL, the assay limit of detection, were reported by the software as “>10 IU/mL”.[10] These values were recoded to 10 IU/mL in this analysis.

### Outcome and covariate definition

The primary aim of this analysis was to describe the distribution of quantitative IGRA responses and their association with participant characteristics. For each participant, TB1 and TB2 responses were calculated by subtracting the nil tube value from the corresponding antigen tube value, resulting, in some cases, in negative values. Nil-subtracted values are hereafter referred to simply as TB1 and TB2 responses.

Two outcomes were defined. First, continuous TB1 and TB2 responses. Second, a binary “TB2-TB1 > 0.6 IU/mL” differential, calculated as TB2 minus TB1, and using a threshold (>0.6 IU/mL) previously proposed as indicative of recent Mtb exposure.[13]

For regression modelling of response magnitude, TB1 and TB2 values were pre-processed to accommodate hurdle models, which require non-negative outcomes. In the primary analysis, values below the manufacturer’s positivity threshold (<0.35 IU/mL) were recoded to 0 IU/mL. All other analyses used the original nil-subtracted values. Tube response-specific qualitative results were derived from quantitative values according to the manufacturer’s criteria.[10] TB1 and TB2 responses were classified as positive if ≥0.35 IU/mL and ≥25% of the nil value, negative if <0.35 IU/mL and ≥25% of the nil, and indeterminate if nil was high (>8 IU/mL, or the antigen response was insufficiently low (<25% of the nil) in the presence of a low nil-subtracted mitogen response (<0.5 IU/mL).[10] To assess the implications of the possibility that responses below the manufacturer’s threshold may still reflect true Mtb exposure in general populations, and consequences for Mtb immunoreactivity prevalence estimates and inference about age- and sex-specific transmission risks, hypothetical alternative thresholds for defining positive test results were explored.

Sociodemographic and clinical factors known to influence or correlate with immune Mtb responses or TB risk were obtained from the primary study. These included age (years), sex (male/female), and self-reported HIV status (positive/negative), previous TB treatment (yes/no) and previous household TB contact (yes/no). Household TB contact was defined as having lived in the same household as a person treated for TB.

### Statistical analysis

Participant characteristics and TB1 and TB2 values were summarised using counts and proportions, means (± standard deviation), or medians (interquartile range). Distributions were visualised using boxplots and histograms.

Tube response agreement was assessed using the residual correlation parameter (ρ) from joint multivariate normal models of TB1 and TB2 values. To characterise the distributions of TB1 and TB2 responses and their associations with age and sex, and other individual covariates, two regression approaches were used.

First, hurdle lognormal models were fitted to estimate 1) the probability that a response fell below the manufacturer’s positivity threshold (<0.35 IU/mL), i.e. the hurdle component, which captured responses equivalent to “0 IU/mL” and did not contribute to the continuous response distribution, and 2) the mean and standard deviation of log-transformed responses ≥0.35 IU/mL, i.e. the lognormal component, which captured the distribution of continuous responses, conditional on clearing the hurdle, and an upper bound at 10 IU/mL. Second, the odds of a TB2-TB1 differential exceeding 0.6 IU/mL were estimated using logistic regression. For each outcome, models were adjusted for age and sex, specified using a thin plate regression spline with 3 basis functions (models with more basis functions performed similarly). The hurdle probability of the hurdle lognormal was also specified as a spline function of age and sex, as above. All other covariates were specified as binary variables, added to the age- and sex-adjusted models.

To assess the implications of hypothetical alternative test positivity thresholds on inference about Mtb immunoreactivity and transmission, qualitative results were reclassified over a range of hypothetical positivity thresholds from 0 to 0.5 IU/mL, in 0.1 IU/mL intervals (higher thresholds are not reported due to progressively fewer participants with very high responses, thus unstable estimates). Across thresholds, age- and sex-specific prevalence of immunoreactivity was estimated using logistic regression models, with positivity as the outcome, excluding indeterminate results. Age and sex were specified as a spline function, as above. Implied sex-specific age trends in transmission risk were inferred from the annual change in predicted immunoreactivity probability, as per Middelkoop et al[23], calculated across the range of thresholds considered.

Models were fitted in the R environment for statistical computing v4.3.3[24] using the brms package v2.2.0, an interface for probabilistic parameter sampling via Hamiltonian Monte Carlo using the No U-Turn Sampler in Stan.[25] Weakly informative priors were specified for intercepts, standard deviations and regression coefficients. Models were fitted using Markov chain Monte Carlo with four chains, 2,000 iterations per chain, and a target acceptance probability of 0.999. Posterior means and 95% percentile intervals were calculated from 4,000 post-warm-up draws. Convergence and sampling efficiency were evaluated using trace plots, effective sample sizes, and the Gelman-Rubin potential scale reduction statistic. Model fit was assessed through posterior predictive distribution plots. Alternative covariate sets and model specifications were compared using leave-one-out cross-validation, as implemented in the loo package.[26] Complete case analysis was used. Full R analysis code is available from https://github.com/mphadsphiri/Quantitative-IGRA.

### Ethics

The study was approved by the Kamuzu University of Health Sciences Research Ethics Committee (COMREC, Ref P.04/22/3611), and the London School of Hygiene & Tropical Medicine Research Ethics Committee (Ref 2774). Written informed consent was obtained for all participants. Participants age less than 18 years provided written assent in addition to written informed consent from a guardian or legally acceptable representative. Illiterate participants or guardians provided a thumbprint, countersigned by an impartial witness. Participants with positive test results were managed according to national guidelines.[27] No treatment was provided as part of the study; however participants received counselling on TB symptoms and appropriate health-seeking behaviour at enrolment and when results were communicated. Participants living with HIV were advised to seek TB preventive therapy if they had not previously received it.

## Results

### Study participant characteristics

Overall, 2,895 participants with QuantiFERON-TB Gold Plus (QFT-Plus) results were included in the analysis. Of these, 40.0% were male, and 43.4% were aged 10-19 years. The median age was 21 years (interquartile range: 16-28 years). Overall, 8.7% (181/2,091) of participants reported having HIV, nearly all (97.2% [176/181]) of whom reported taking antiretroviral therapy. Few (1.1% [31/2,895]) participants reported fever; 1.2% (34/2,895) reported cough with duration ≥ 2 weeks. Overall, 17.4% (503/2,895) of participants had QFT-Plus positive results and 2.1% (62/2,895) had indeterminate results. (Table 1)

**Table 1:**
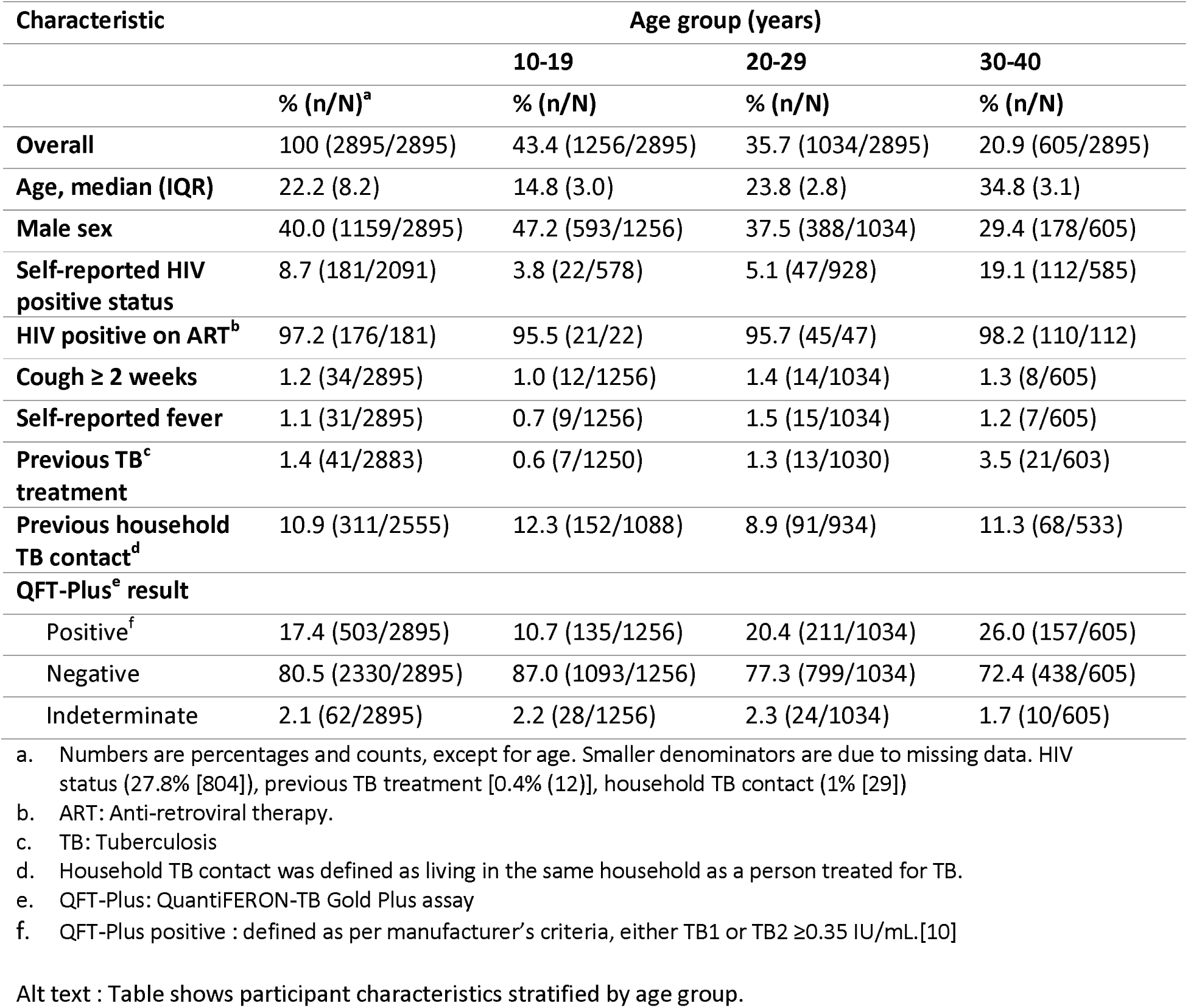
Participant characteristics and QuantiFERON-TB Gold Plus test results by age.

### TB1 and TB2 response-specific QFT-Plus qualitative results

Qualitative TB1 and TB2 results were similar. 13.6% (395/2895) of participants had a positive result on both TB1 and TB2. (Table 4) Few participants had discordant TB1 and TB2 results: 1.6% (47/2895) were positive on TB1 but negative on TB2, and 2.0% (59/2895) were negative on TB1 but positive on TB2. (Table 4) Indeterminate results occurred in 2.1% (62/2895) of participants, nearly all (96.9% [62/64]) on both TB1 and TB2 tubes. One participant had a positive result on TB1 and an indeterminate result on TB2, and another vice versa. (Table 4)

### Distribution of continuous TB1 and TB2 responses

Quantitative TB1 and TB2 interferon gamma (IFN-γ) responses were highly positively-skewed. (Figure 1 and Supplementary Figure 1) Overall, the mean and median TB1 responses were 0.41 IU/mL (±standard deviation: 1.52 IU/mL) and 0.01 (interquartile range [IQR]: -0.01-0.06) IU/mL, respectively. The corresponding values for TB2 were mean 0.43 (±1.53) IU/mL and median 0.01 (IQR: 0.00-0.07) IU/mL. Few participants (1.7% [48/2895] for TB1 and 0.4% [13/2895] for TB2) had responses which exceeded 10 IU/mL; these were recoded to 10 IU/mL. (Supplementary Table 1)

**Figure 1:**
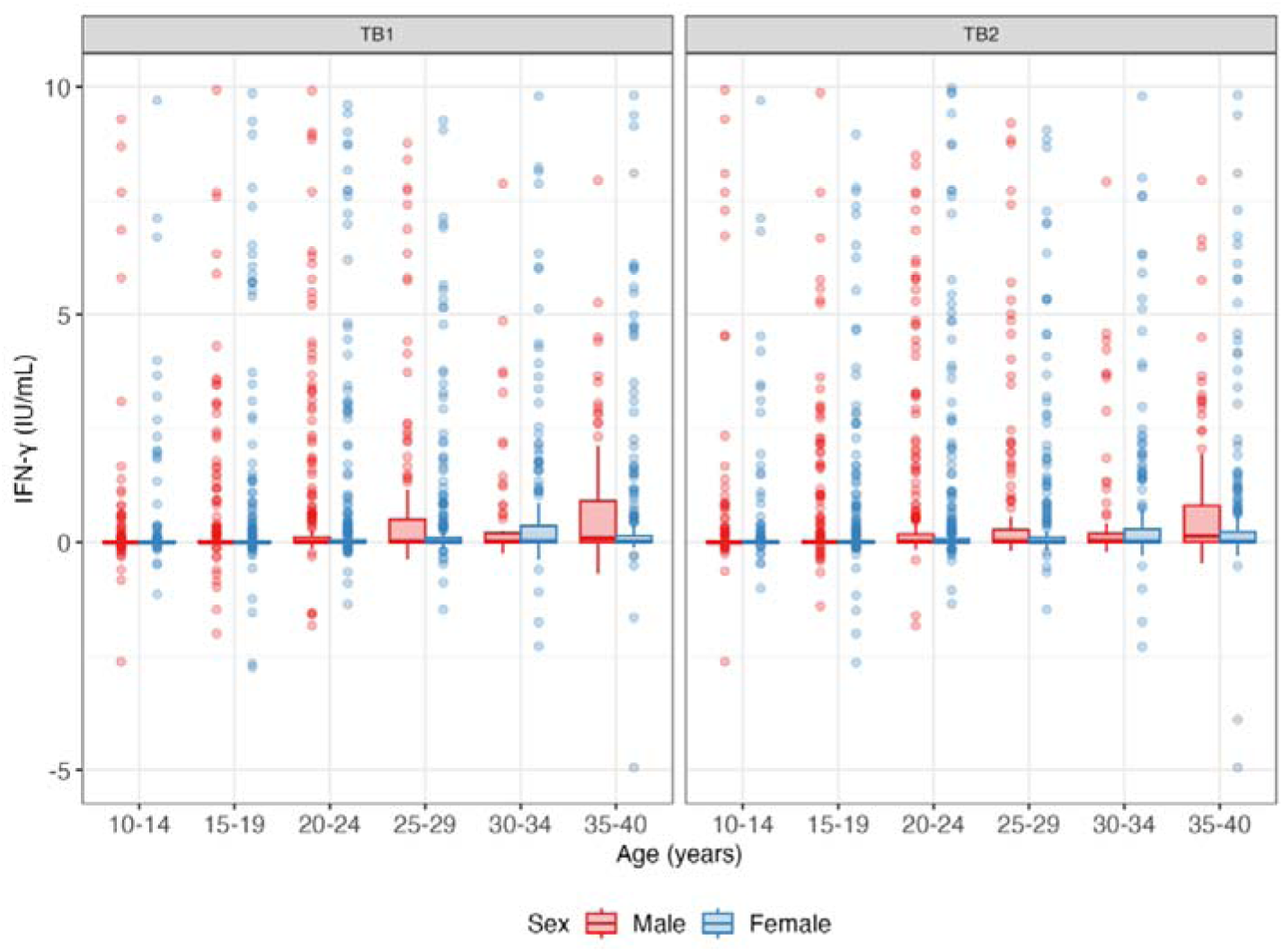
Boxplot showing the distributions of nil-subtracted TB1 and TB2 values by age and sex. IFN-γ: interferon gamma. IU/mL: international units per millilitre. Note: IFN-γ values are right-truncated at the assay upper limit of detection (10 IU/mL). 12 values less than -5 IU/mL are not shown for clarity. Alt text: Figure shows distribution of quantitative interferon-gamma responses (QuantiFERON-TB Gold Plus TB1 and TB2) by age and sex.

### Factors associated with QuantiFERON-TB Gold Plus TB1 and TB2 response magnitude

The distributions of TB1 and TB2 responses above the manufacturer’s threshold showed no consistent pattern across the age range, and were similar by sex. (Figure 2)

**Figure 2:**
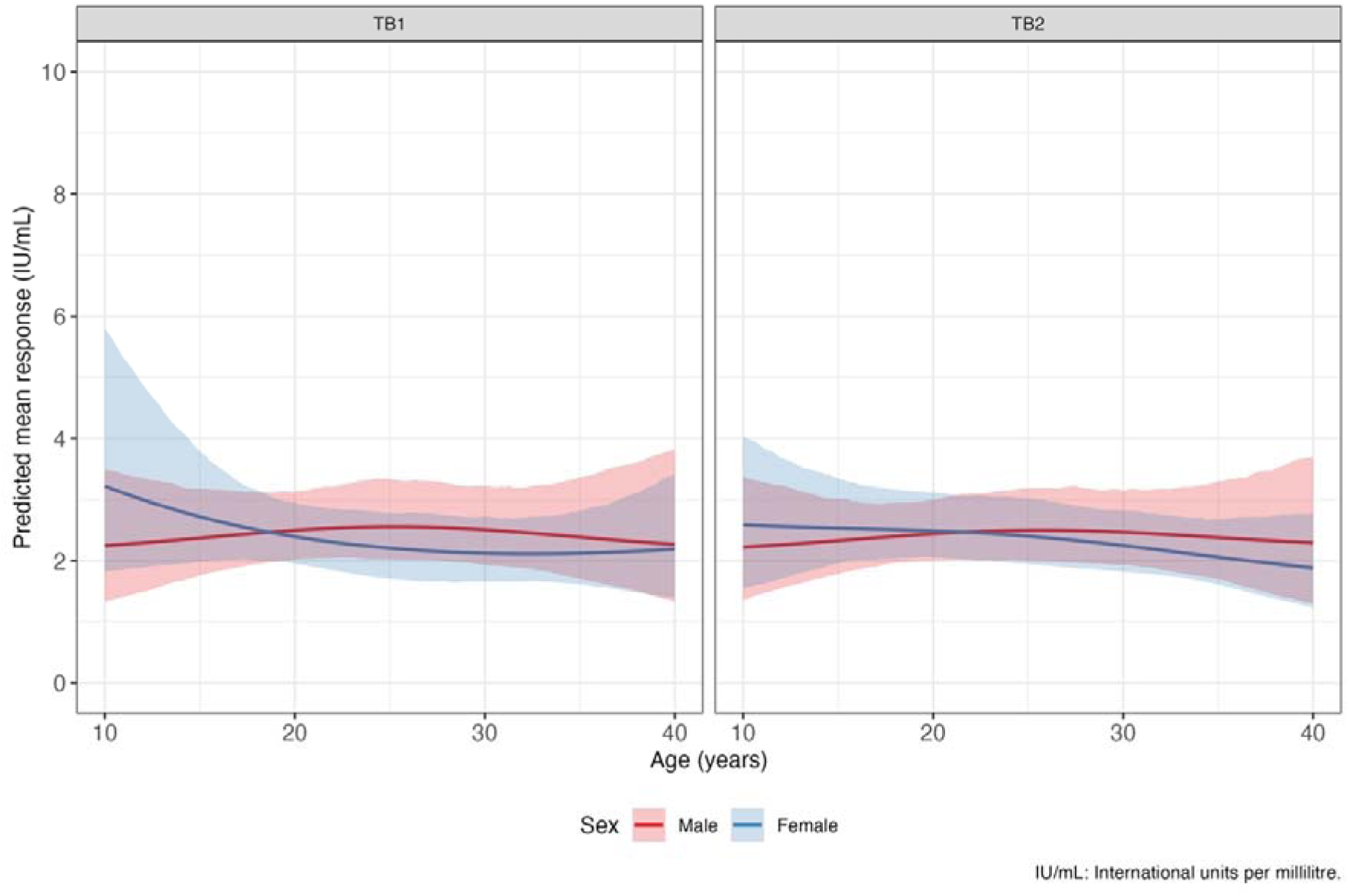
Predicted mean age- and sex-specific QuantiFERON-TB Gold Plus (QFT-Plus) responses by antigen tube among participants with a positive QFT-Plus result. Responses were predicted from the lognormal components on Bayesian hurdle lognormal regression models for nil-subtracted TB1 and TB2 responses adjusted for age and sex. Age and sex were specified as a thin plate spline with 3 basis functions. Curves are posterior means, and shaded areas are 95% credible intervals. Posterior predicted distributions for the hurdle components of the models are shown in Supplementary Figure 2. Alt text: Figure shows predicted mean interferon-gamma responses by tube (QuantiFERON-TB Gold Plus [QFT-Plus] TB1 and TB2) among participants with a positive QFT-Plus result (TB1 or TB2 ≥0.35 IU/mL).

There was no evidence that HIV status, previous TB treatment, or household TB contact history were associated with TB1 or TB2 response magnitude (Table 2). Associations between participant characteristics and TB1 and TB2 response magnitude estimated from the lognormal components of hurdle lognormal models, are summarised in Table 3.Posterior predictive distributions showed that the models reproduced observed age- and sex-specific proportions of responses below 0.35 IU/mL. The probability of a having response ≥0.35 IU/mL increased with age, diverging by sex from late adolescence. (Supplementary Figure 2)

**Table 2:**
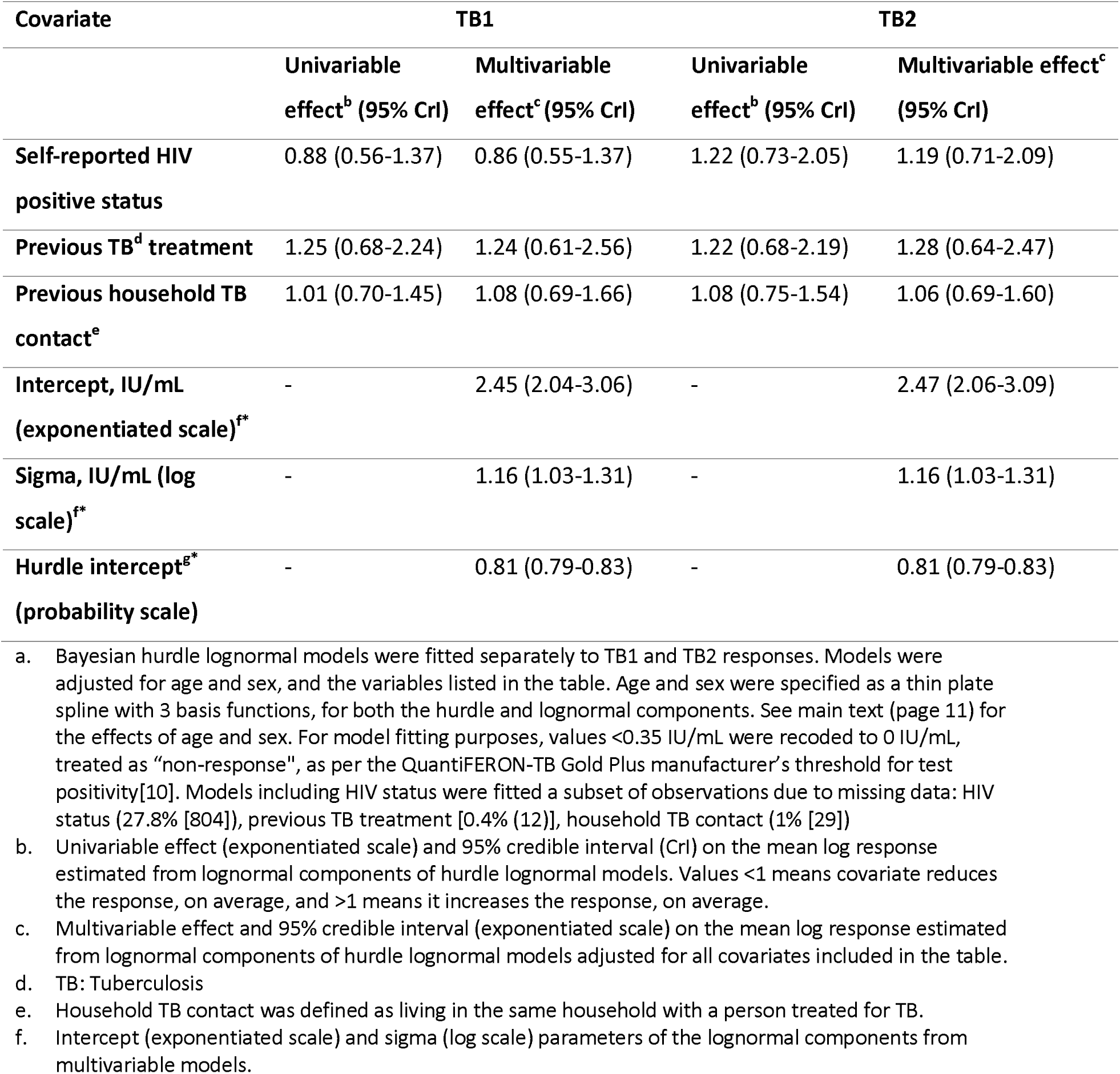
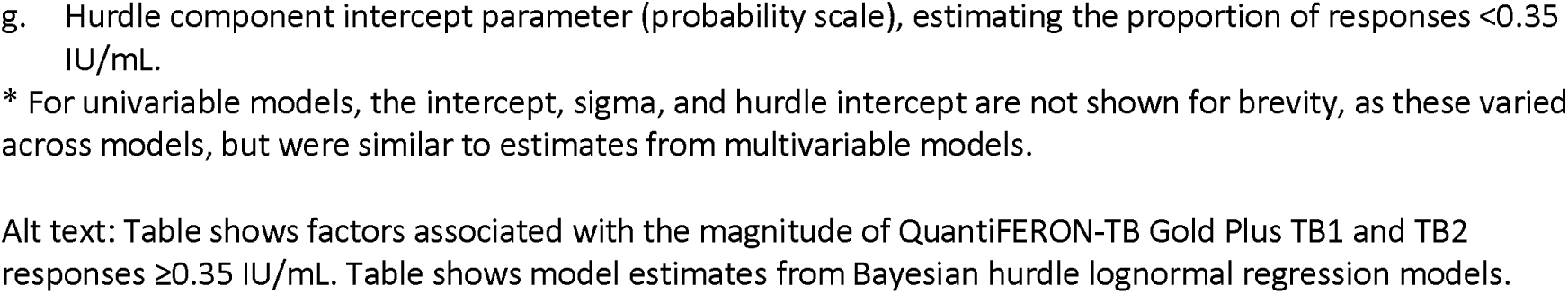
Factors associated with the magnitude of QuantiFERON-TB Gold Plus TB1 and TB2 responses ≥0.35 IU/mL, estimated from Bayesian hurdle lognormal regression models adjusted for age and sex^a^.

| Covariate | TB1 |  | TB2 |  |
| --- | --- | --- | --- | --- |
|  | Univariable effect<sup>b</sup> (95% CrI) | Multivariable effect<sup>c</sup> (95% CrI) | Univariable effect<sup>b</sup> (95% CrI) | Multivariable effect<sup>c</sup> (95% CrI) |
| **Self-reported HIV positive status** | 0.88 (0.56-1.37) | 0.86 (0.55-1.37) | 1.22 (0.73-2.05) | 1.19 (0.71-2.09) |
| **Previous TB<sup>d</sup> treatment** | 1.25 (0.68-2.24) | 1.24 (0.61-2.56) | 1.22 (0.68-2.19) | 1.28 (0.64-2.47) |
| **Previous household TB contact<sup>e</sup>** | 1.01 (0.70-1.45) | 1.08 (0.69-1.66) | 1.08 (0.75-1.54) | 1.06 (0.69-1.60) |
| **Intercept, IU/mL (exponentiated scale)<sup>f\*</sup>** | - | 2.45 (2.04-3.06) | - | 2.47 (2.06-3.09) |
| **Sigma, IU/mL (log scale)<sup>f\*</sup>** | - | 1.16 (1.03-1.31) | - | 1.16 (1.03-1.31) |
| **Hurdle intercept<sup>g\*</sup> (probability scale)** | - | 0.81 (0.79-0.83) | - | 0.81 (0.79-0.83) |

- Bayesian hurdle lognormal models were fitted separately to TB1 and TB2 responses. Models were adjusted for age and sex, and the variables listed in the table. Age and sex were specified as a thin plate spline with 3 basis functions, for both the hurdle and lognormal components. See main text (page 11) for the effects of age and sex. For model fitting purposes, values $< 0.35$ IU/mL were recoded to 0 IU/mL, treated as "non-response", as per the QuantiFERON-TB Gold Plus manufacturer's threshold for test positivity [10]. Models including HIV status were fitted a subset of observations due to missing data: HIV status (27.8% [804]), previous TB treatment [0.4% (12)], household TB contact (1% [29]) - Univariable effect (exponentiated scale) and 95% credible interval (CrI) on the mean log response estimated from lognormal components of hurdle lognormal models. Values $< 1$ means covariate reduces the response, on average, and $> 1$ means it increases the response, on average. - Multivariable effect and 95% credible interval (exponentiated scale) on the mean log response estimated from lognormal components of hurdle lognormal models adjusted for all covariates included in the table. - TB: Tuberculosis - Household TB contact was defined as living in the same household with a person treated for TB. - Intercept (exponentiated scale) and sigma (log scale) parameters of the lognormal components from multivariable models.
g. Hurdle component intercept parameter (probability scale), estimating the proportion of responses <0.35 IU/mL.
\* For univariable models, the intercept, sigma, and hurdle intercept are not shown for brevity, as these varied across models, but were similar to estimates from multivariable models.
Alt text: Table shows factors associated with the magnitude of QuantiFERON-TB Gold Plus TB1 and TB2 responses $\geq 0.35$ IU/mL. Table shows model estimates from Bayesian hurdle lognormal regression models.

**Table 3:**
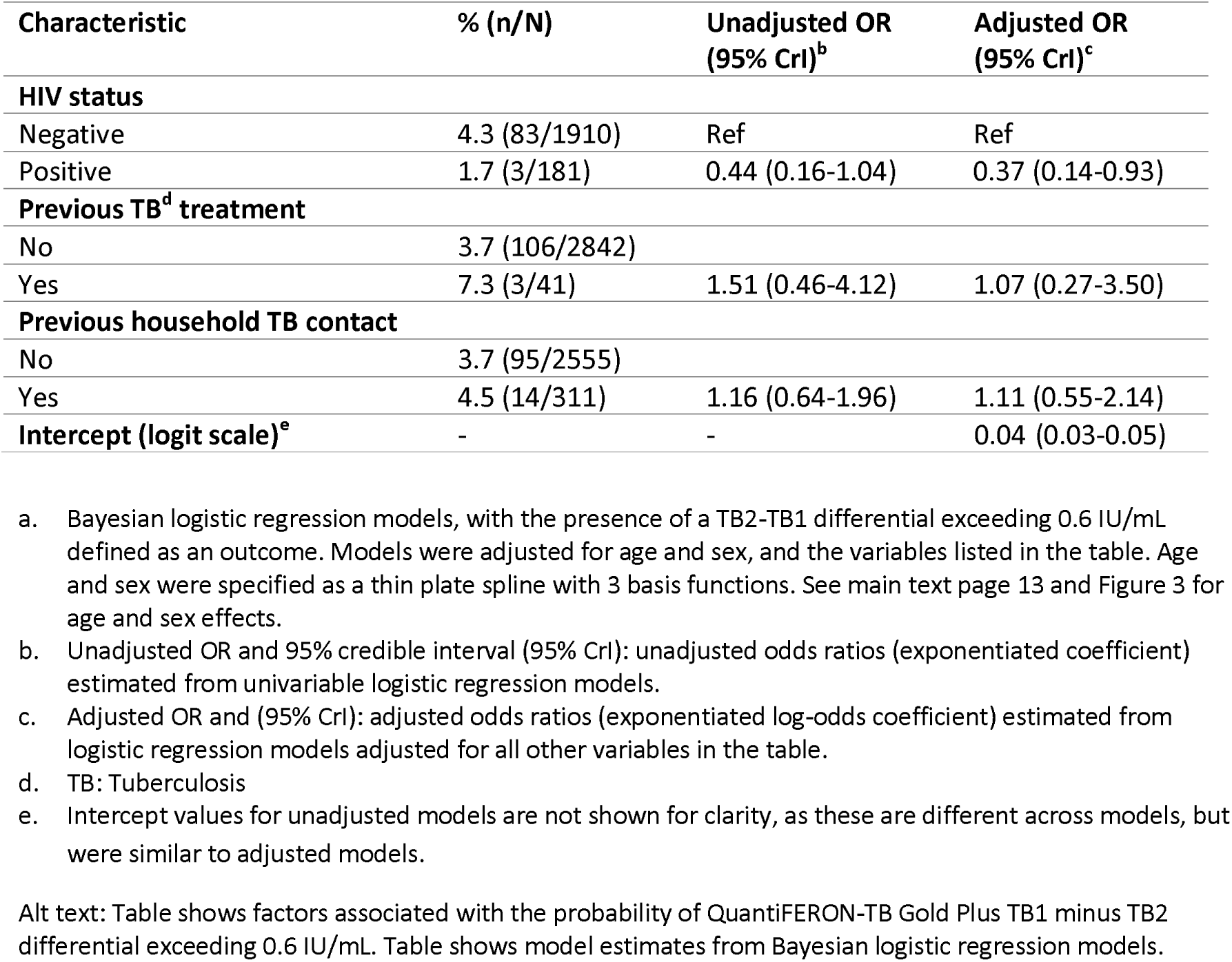
Factors associated with a QuantiFERON-TB Gold Plus TB2-TB1 differential exceeding 0.6 IU/mL, estimated from Bayesian logistic regression models adjusted for age and sex^a^.

| Characteristic | % (n/N) | Unadjusted OR (95% CrI)<sup>b</sup> | Adjusted OR (95% CrI)<sup>c</sup> |
| --- | --- | --- | --- |
| **HIV status** |  |  |  |
| Negative | 4.3 (83/1910) | Ref | Ref |
| Positive | 1.7 (3/181) | 0.44 (0.16-1.04) | 0.37 (0.14-0.93) |
| **Previous TB<sup>d</sup> treatment** |  |  |  |
| No | 3.7 (106/2842) |  |  |
| Yes | 7.3 (3/41) | 1.51 (0.46-4.12) | 1.07 (0.27-3.50) |
| **Previous household TB contact** |  |  |  |
| No | 3.7 (95/2555) |  |  |
| Yes | 4.5 (14/311) | 1.16 (0.64-1.96) | 1.11 (0.55-2.14) |
| **Intercept (logit scale)<sup>e</sup>** | - | - | 0.04 (0.03-0.05) |

Alt text: Table shows factors associated with the probability of QuantiFERON-TB Gold Plus TB1 minus TB2 differential exceeding 0.6 IU/mL. Table shows model estimates from Bayesian logistic regression models.

**Table 4:** Tube-specific response QuantiFERON-TB Gold Plus results, defined according to the manufacturer’s criteria, across participant characteristics.

| Characteristic | TB1 & TB2 positive, % (n/N) | TB1 & TB2 negative, % (n/N) | TB1 positive & TB2 negative, % (n/N) | TB1 negative & TB2 positive, % (n/N) | TB1 & TB2 indeterminate, % (n/N) | TB1 positive & TB2 indeterminate, % (n/N) | TB1 indeterminate & TB2 positive, % (n/N) |
| --- | --- | --- | --- | --- | --- | --- | --- |
| Overall | 13.6 (395/2895) | 80.5 (2330/2895) | 1.6 (47/2895) | 2.0 (59/2895) | 2.1 (62/2895) | <0.1 (1/2895) | <0.1 (1/2895) |
| **Age** |  |  |  |  |  |  |  |
| 10-19 | 8.0 (100/1256) | 87.0 (1093/1256) | 0.7 (9/1256) | 1.9 (24/1256) | 2.2 (28/1256) | 0.1 (1/1256) | 0.1 (1/1256) |
| 20-29 | 16.0 (165/1034) | 77.3 (799/1034) | 2.5 (26/1034) | 1.9 (20/1034) | 2.3 (24/1034) | 0.0 (0/1034) | 0.0 (0/1034) |
| 30-40 | 21.5 (130/605) | 72.4 (438/605) | 2.0 (12/605) | 2.5 (15/605) | 1.7 (10/605) | 0.0 (0/605) | 0.0 (0/605) |
| **Sex** |  |  |  |  |  |  |  |
| Male | 14.3 (166/1159) | 79.0 (916/1159) | 1.9 (22/1159) | 2.3 (27/1159) | 2.2 (26/1159) | 0.1 (1/1159) | 0.1 (1/1159) |
| Female | 13.2 (229/1736) | 81.5 (1414/1736) | 1.4 (25/1736) | 1.8 (32/1736) | 2.1 (36/1736) | 0.0 (0/1736) | 0.0 (0/1736) |

a. QuantiFERON-TB Gold Plus manufacturer's criteria for positivity, i.e. antigen response $\geq 0.35$ IU/mL.[10] No participant had negative TB1 and indeterminate TB2 results, or vice versa.
Alt text: Table shows qualitative TB1- and TB2-specific QuantiFERON-TB Gold Plus results overall, and by age and sex.

### Correlation between TB1 and TB2 responses

Correlation between TB1 and TB2 responses was high: posterior mean residual correlation parameter: 0.86 (95% CrI: 0.85-0.87). Correlation increased weakly with age and was slightly lower in males than in females. (Supplementary Table 2)

### TB2-TB1 differential

The mean TB2 minus TB1 (TB2-TB1) differential was 0.02 (±0.81) IU/mL (Supplementary Figure 3). No factors were associated with TB2-TB1. Overall, 3.8% (109/2895) of participants had a TB2-TB1 differential exceeding 0.6 IU/mL. (Table 3). The odds of an excess TB2 response showed a weak age trend. Figure 3 shows corresponding trends in predicted probabilities of an excess TB2 response by age and sex. Before age 20 years, the trend in females was, on average, higher than males. However, from late adolescence, the trend in males increased steadily, exceeding that in females from the early 20s through to 40 years. However, 95% CrIs overlapped.

**Figure 3:**
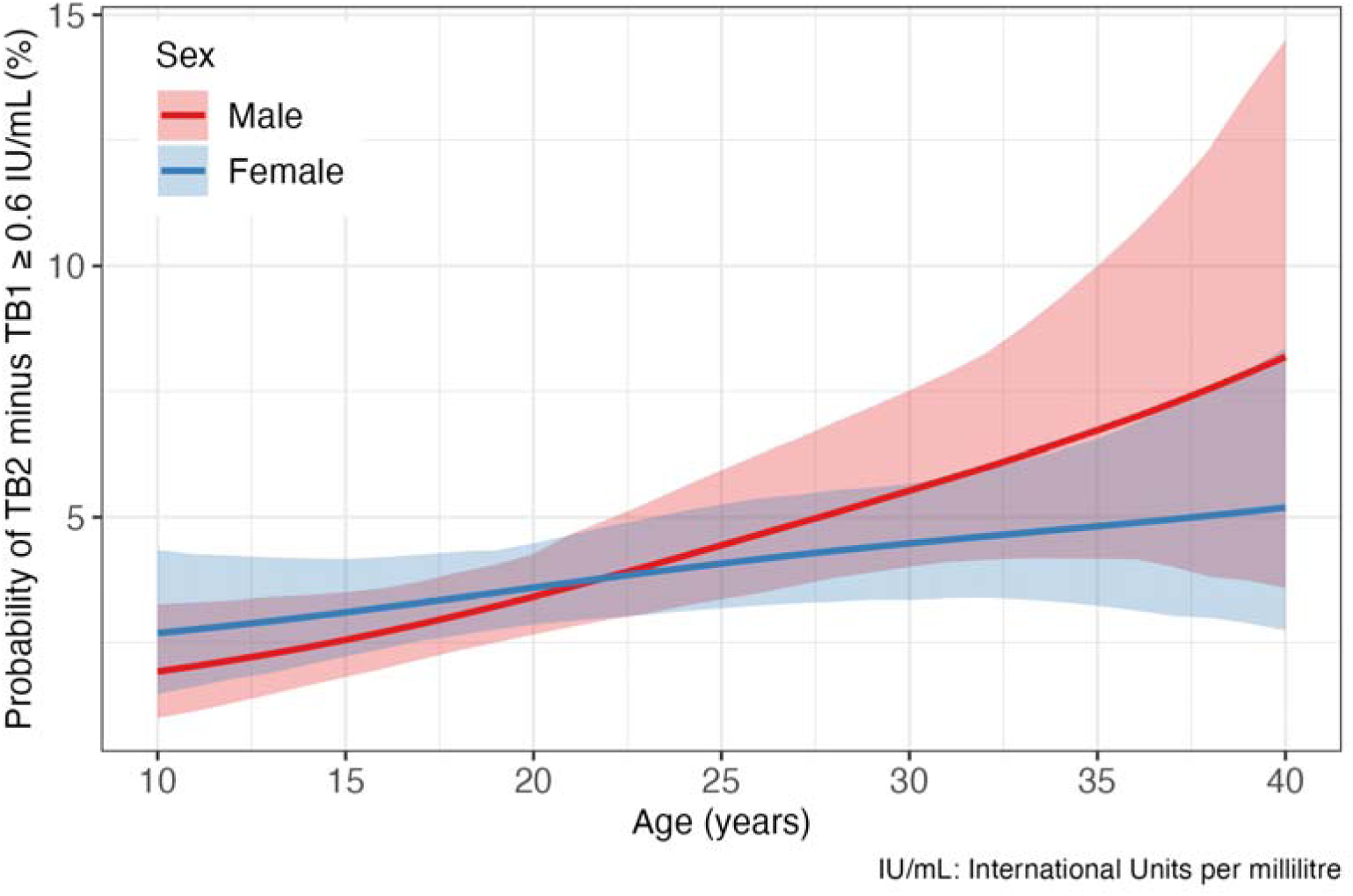
Age- and sex-specific predicted probability of a QuantiFERON-TB Gold Plus TB2 minus TB1 differential exceeding 0.6 international units per millilitre. A TB2-TB1 differential has been previously reported to be associated with recent *Mycobacterium tuberculosis* exposure. Probabilities are exponentiated odds from a Bayesian logistic regression model adjusted for age and sex. Age was specified as a thin-plate spine with 3 basis functions for each sex. Curves are posterior means, with shaded regions representing associated 95% credible intervals. Alt text: Figure shows model predicted probability of a QuantiFERON-TB Gold Plus TB2 minus TB1 differential exceeding 0.6 IU/mL by age and sex.

In contrast, the odds of an excess TB2 response were reduced in those living with HIV (OR: xxx). (Table 3) However, there was no evidence for associations with previous TB treatment or household TB contact.

### Implications of hypothetical alternative test positivity thresholds on inference about transmission

When QFT-Plus positivity was recalculated at hypothetical alternative thresholds, an additional 100 participants were classified as positive at ≥0.2 IU/mL and a further 129 at ≥0.1 IU/mL, corresponding to 3.5% and 8.0% absolute percent point increases in positivity, respectively, compared to the QFT-Plus threshold (17.8% [503/2895] at ≥0.35 IU/mL).

As expected, predicted estimates of Mtb immunoreactivity prevalence increased at lower thresholds. (Figure 4) Corresponding annual risk of immunoreactivity conversion estimates reached a maximum predicted mean of 3.3% (0.6%-5.8%) in 40-year-old males at 0.1 IU/mL cut-off. (Figure 5) At higher thresholds, predicted age- and sex-specific immunoreactivity prevalence increased faster among males, and corresponding male-to-female ratios suggested that male prevalence exceeded female prevalence at earlier ages in adolescence than at lower thresholds. At 19 years, predicted male-to-female (M:F) immunoreactivity prevalence ratios were 0.90 (0.83-0.99) and 1.02 (0.89-1.15) at 0.1 IU/mL and 0.5 IU/mL thresholds, respectively. The M:F prevalence ratio at the QFT-Plus manufacturer threshold (0.35 IU/mL) was 0.99 (0.88-1.11). However, 95% CrIs for M:F ratios included 1, i.e. prevalence in males was equal to that in females. (Figure 6)

**Figure 4:**
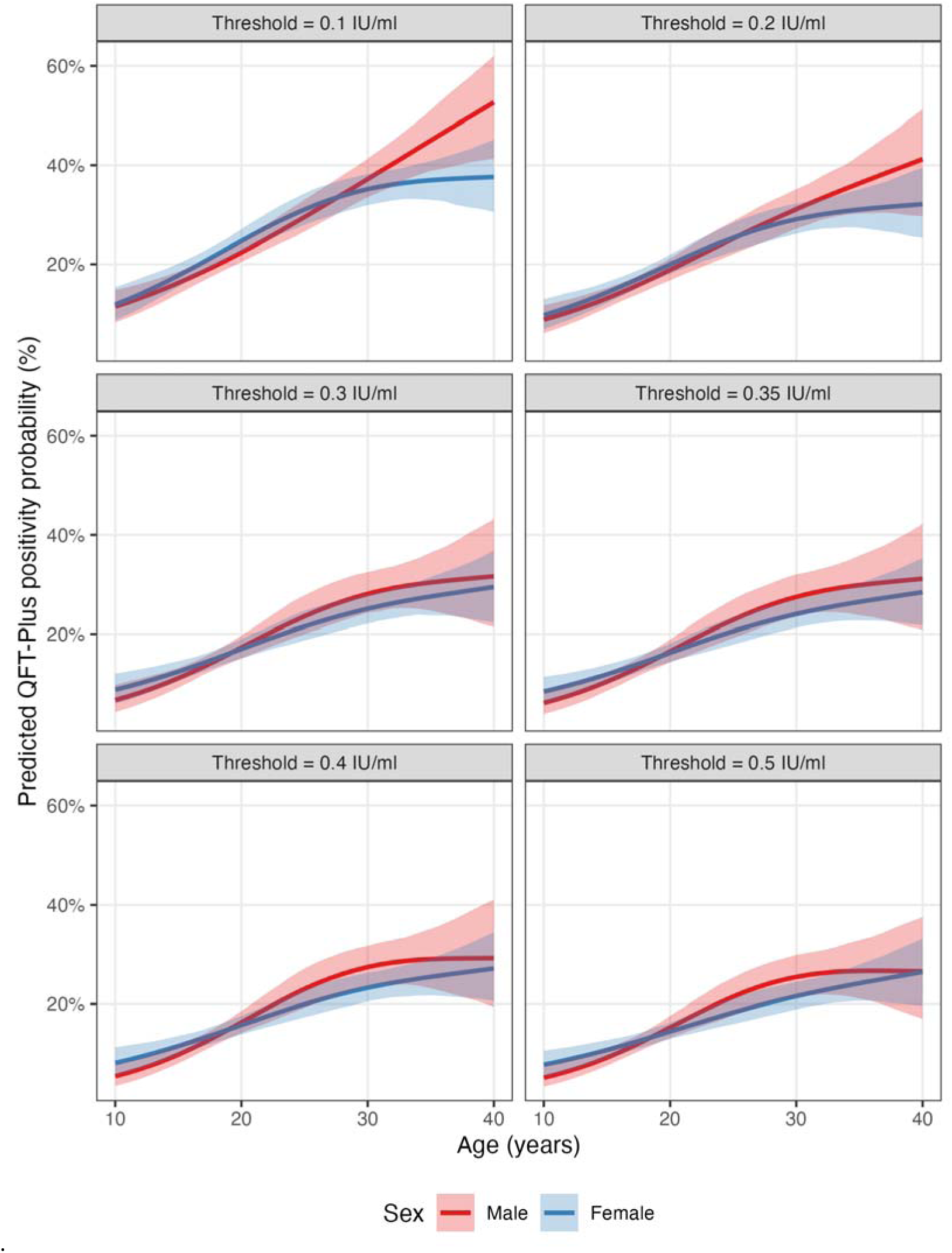
Age- and sex-specific probability of *Mycobacterium tuberculosis* (Mtb) immunoreactivity at hypothetical alternative interferon gamma level thresholds for defining test positivity. Probabilities are computed as exponentiated odds predicted by a Bayesian logistic regression model, adjusted for age and sex. Age was specified as a spline function with a sex-specific offset. Mtb immunoreactivity was defined as a positive QuantiFERON-TB Gold Plus test result. Curves are posterior means, with shaded regions representing associated 95% credible intervals. Alt text: Age- and sex-specific probability of *Mycobacterium tuberculosis* (Mtb) immunoreactivity across hypothetical alternative interferon gamma level thresholds for defining test positivity, including the QFT-Plus test manufacturer’s threshold (≥0.35 IU/mL).

**Figure 5:**
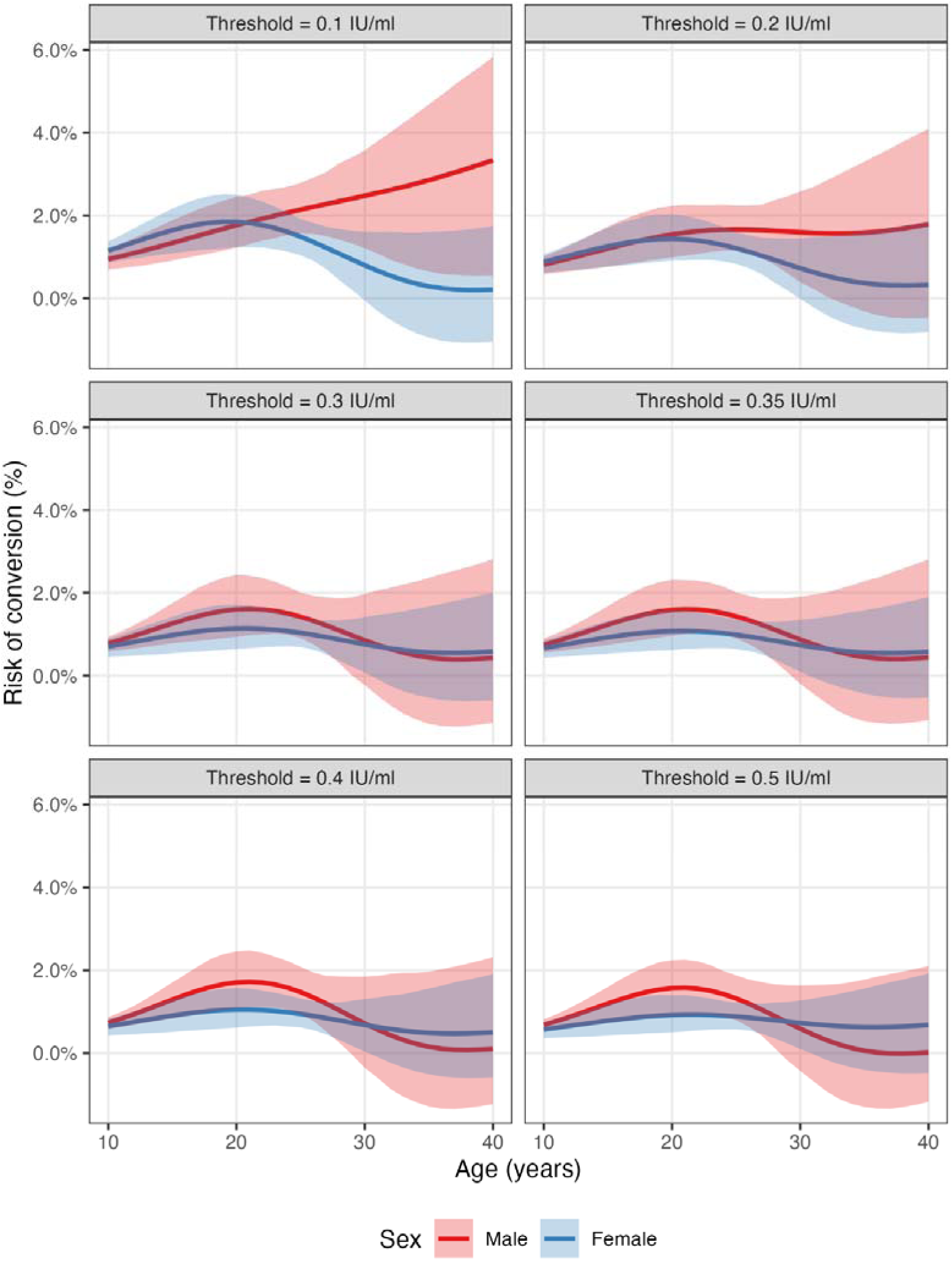
Age and sex specific annual risk of *Mycobacterium tuberculosis* conversion by hypothetical alternative thresholds interferon gamma level cut-offs for positivity. Annual risk of conversion was calculated from the annual change in prevalence estimates, divided by prevalence at the previous age, as per Middelkoop et al[23] Prevalence was obtained by exponentiating the odds predicted by a Bayesian logistic regression model adjusted for age and sex. Age was specified as a thin-plate spline with 3 basis functions. Mtb immunoreactivity was defined as a positive QuantiFERON-TB Gold Plus test result. Curves are posterior means, with shaded regions representing associated 95% credible intervals. Alt text: Age- and sex-specific probability of annual risk of *Mycobacterium tuberculosis* (Mtb) immunoreactivity conversion across hypothetical alternative interferon gamma level thresholds for defining test positivity, including the QFT-Plus test manufacturer’s threshold (≥0.35 IU/mL).

**Figure 6:**
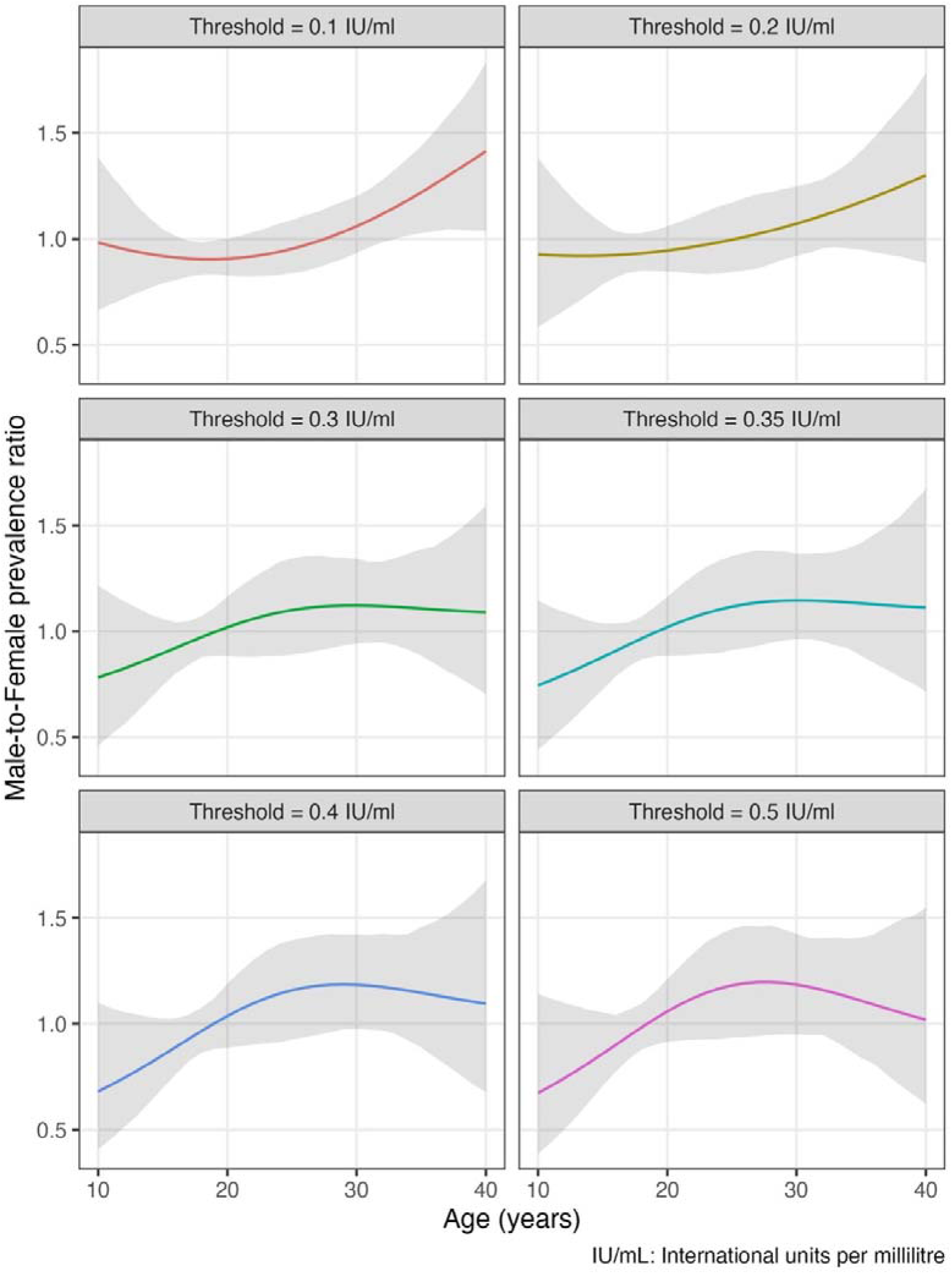
Age-specific male-to-female ratios of *Mycobacterium tuberculosis* (Mtb) immunoreactivity prevalence across interferon gamma level thresholds for defining test positivity. Mtb immunoreactivity prevalence estimates were obtained by exponentiating the odds predicted from Bayesian logistic regression models adjusted for age and sex. Age was specified as a thin plate spine with 3 basis functions. Mtb immunoreactivity was defined as a positive QuantiFERON-TB Gold Plus test result. QFT-Plus test manufacturer threshold is 0.35 international units per millilitre. Other thresholds are hypothetical. Curves are posterior means, with shaded regions representing associated 95% credible intervals. Alt text: Age-specific male-to-female ratios of *Mycobacterium tuberculosis* (Mtb) immunoreactivity prevalence across hypothetical alternative interferon gamma level thresholds for defining test positivity, including the QFT-Plus test manufacturer’s threshold (≥0.35 IU/mL).

## Discussion

This study characterised the magnitude and distribution of interferon gamma responses to *Mycobacterium tuberculosis* (Mtb), measured by QuantiFERON-TB Gold Plus (QFT-Plus), among 10–40-year-olds in high-burden communities in Blantyre, Malawi. As expected, the probability of tube-specific QFT-Plus results varied by age and diverged by sex from late adolescence. However, conditional TB1 and TB2 response magnitudes were similar across age and between sexes. A TB2 excess response, proposed as a marker of recent Mtb infection, was associated HIV-positive status. At higher hypothetical thresholds for test positivity, sex-specific age trends in prevalence and annual risk of immunoreactivity diverged earlier during adolescence. In high burden settings, quantitative IGRA responses could provide insights into transmission dynamics and guide targeted strategies to reduce Mtb exposure risk and disease progression.

We observed Mtb-specific interferon gamma (IFN-γ) response distributions as expected in populations with heterogeneous exposure risk. In Karonga, northern Malawi, community-recruited individuals showed predominantly low responses to ESAT-6 and MPT-64, with few high responders.[28] However, Karonga’s rural TB epidemiology differs from Blantyre’s urban epidemic, and the in-house assay used in Karonga contained different antigens from QFT-Plus, limiting direct comparison.[21,29,30] Most published studies reporting quantitative IGRA responses have enrolled individuals selected for Mtb exposure through contact investigations, clinical cohorts, or test performance studies, predominantly in low- to moderate-incidence countries.[7] This study contributes population-level data from randomly sampled individuals in a high-incidence setting where such data remain sparse.

Age-related differences in Mtb IFN-γ responses may arise through multiple mechanisms. Maturation and the development of antigen-specific T-cell memory across childhood and adolescence influence the capacity to mount detectable responses.[31,32]. Cumulative Mtb exposure increases with age, reflecting more opportunities for high TB risk contacts.[19] In our study, however, whereas age- and sex-were associated with the probability of being positive at the manufacturer’s threshold positivity, these factors were not associated with the magnitude of response.. This suggests that cumulative exposure and immune priming determine whether a response is elicited, rather than amplifying response magnitude once immunoreactivity is established. These mechanisms are not mutually exclusive.

In animal models, sex hormones modulate Mtb IFN-γ production, with testosterone suppressing responses.[33] In this study, however, we found little evidence that the magnitude of IFN-γ responses differed by sex across age. Clarifying the immunological mechanisms underlying sex differences in Mtb responses and their relationship to infection and disease risk requires longitudinal and experimental studies that examine biological mechanisms beyond epidemiological associations. Such evidence would inform inference from prevalence surveys and clinical trials, and augment ongoing efforts to address sex disparities in TB.

IGRA response magnitude did not differ meaningfully by HIV status. Nearly all participants with HIV reported taking antiretroviral therapy and thus likely had reconstituted immunity sufficient to mount Mtb-specific responses. Similarly, household TB contact and previous TB treatment, established correlates of IGRA positivity, and previously reported in the original study,[18] were not associated with response strength. The study likely lacked precision to detect these known associations.

A small proportion of individuals had a TB2-TB1 differential exceeding 0.6 IU/mL, the threshold proposed to identify recent exposure. Evidence for this marker remains inconsistent. Four studies in low-incidence settings reported positive associations with recent exposure,[11,13,34,35] while a fifth did not.[36] In high-burden settings, the differential did not discriminate recent from remote infection: among 15-24 year olds in Zambia and South Africa, QFT-Plus conversion over two years was not associated with TB2-TB1 >0.6 IU/mL.[17] In our study, we found no evidence that age, sex, previous TB treatment or household TB contact were associated with the odds of TB2-TB1 exceeding 0.6 IU/mL. However, HIV positivity was associated with reduced odds of a TB2 excess. TB2 responses are known to be diminished by HIV infection. However, without data on timing of exposure or disease outcomes, the relevance of this differential for this population remains unknown. If confirmed, identifying and treating individuals with recent infection, a key driver of incidence, may be possible in community-based IGRA surveys.

Reclassifying test results using hypothetical alternative thresholds revealed expected patterns: lower thresholds increased estimated immunoreactivity prevalence and annual risk of conversion. Notably, at higher thresholds, sex-specific age trends diverged further during adolescence. The manufacturer’s 0.35 IU/mL threshold represents a trade-off between sensitivity and specificity, estimated against TB disease in predominantly low-incidence populations.[7] Nevertheless, it is plausible that responses below this threshold but above zero are true Mtb-specific responses; excluding them from interventions requiring a positive test, such as post-infection vaccines, may be unjust and, if they sustain transmission, counterproductive.

Several limitations merit consideration. First, there is no gold standard for Mtb infection; IGRA responses and associations imperfectly reflect true exposure. However, QFT-Plus sensitivity and specificity, estimated in people with bacteriologically-confirmed TB disease and low risk of Mtb exposure, respectively, exceed 90% and suggest good association with Mtb exposure.[37] Second, the cross-sectional design precludes drawing conclusions about mechanisms underlying the observed associations. Without longitudinal follow-up, the relationship between quantitative responses and incident TB could not be assessed. Third, residual or unmeasured confounders, such as nutritional status, may have influenced the observed associations. HIV status was self-reported. Fourth, granular analyses were limited by small sample sizes, which may have obscured some associations. Fifth, annual risk of conversion estimates ignored reversion. Finally, findings from a single urban setting with research-laboratory blood processing may not generalise to settings with different transmission dynamics or less stringent laboratory conditions. Nevertheless, assay processes followed standard protocols, with support from the manufacturer’s technical team.

In conclusion, population-level variation in quantitative Mtb-specific immune responses clarifies transmission dynamics that fixed positivity thresholds obscure. Quantitative IGRA responses contain epidemiological information about age patterns, sex differences, and exposure gradients that dichotomisation discards. To realise this potential, research must establish how continuous responses correlate with exposure intensity and disease outcomes in general populations in high-burden settings. Positivity thresholds calibrated in low-incidence settings need to be revised to avoid excluding individuals who would benefit from Mtb interventions. Until new tools, such as other cytokine-based or immune-phenotypic assays or RNA-based signatures, become routinely available, IGRAs will remain a key tool for understanding TB epidemiology. Unlocking the public health potential of quantitative IGRA response data could sharpen targeting and impact evaluation of post-infection vaccines, and accelerate progress toward global TB elimination targets.

## Data Availability

All data produced are available online at https://github.com/mphadsphiri/Quantitative–IGRA

## Acknowledgements

We thank study personnel (study participant recruitment and blood sample collection), and Vincent Katunga-Phiri (data management support), at the Malawi Liverpool Wellcome Research Programme; and the Kamuzu University of Health Sciences tuberculosis research laboratory (assay processing); and the Blantyre District Health Office and Malawi National Tuberculosis Programme and Leprosy Elimination Programme (institutional support). Authors also acknowledge support from management, research, research uptake, and programme management teams within The LIGHT Consortium.

Data and R code available at https://github.com/mphadsphiri/Quantitative-IGRA.

## References

1. World Health Organization. Global tuberculosis report 2025 [Internet]. Geneva, Switzerland: World Health Organization; [cited 2025 Nov 25]. p. 76. Report No. Available from: https://www.who.int/publications/i/item/9789240116924

2. Houben RMGJ, Dodd PJ. The Global Burden of Latent Tuberculosis Infection: A Re-estimation Using Mathematical Modelling. PLOS Med. 2016;13:e1002152. doi:10.1371/journal.pmed.1002152

3. Getahun H, Matteelli A, Chaisson RE, Raviglione M. Latent Mycobacterium tuberculosis Infection. N Engl J Med. 2015;372:2127–35. doi:10.1056/NEJMra1405427

4. Schwalb A, Dodd P, Rickman HM, Ugarte-Gil C, Horton KC, Houben RMGJ. Estimating the Global Burden of Viable Mycobacterium Tuberculosis Infection [SSRN Scholarly Paper] [Internet]. Rochester, NY: Social Science Research Network; 2024 [cited 2025 Oct 2]. Available from: https://papers.ssrn.com/abstract=5017943 doi:10.2139/ssrn.5017943

5. Rangaka MX, Cavalcante SC, Marais BJ, Thim S, Martinson NA, Swaminathan S, et al. Controlling the seedbeds of tuberculosis: diagnosis and treatment of tuberculosis infection. The Lancet. 2015;386:2344–53. doi:10.1016/S0140-6736(15)00323-2

6. World Health Organization. WHO consolidated guidelines on tuberculosis: module 3: diagnosis: tests for TB infection [Internet]. Geneva, Switzerland; 2022 [cited 2025 Dec 18]. Report No. Available from: https://www.who.int/publications/i/item/9789240056084

7. Pai M, Denkinger CM, Kik SV, Rangaka MX, Zwerling A, Oxlade O, et al. Gamma Interferon Release Assays for Detection of Mycobacterium tuberculosis Infection. Clin Microbiol Rev. 2014;27:3–20. doi:10.1128/cmr.00034-13

8. World Health Organization. Target product profiles for tuberculosis screening tests [Internet]. World Health Organization; 2025 [cited 2025 Aug 11]. Available from: https://www.who.int/publications/i/item/9789240113572

9. Cobelens F, Suri RK, Helinski M, Makanga M, Weinberg AL, Schaffmeister B, et al. Accelerating research and development of new vaccines against tuberculosis: a global roadmap. Lancet Infect Dis. 2022;22:e108–20. doi:10.1016/S1473-3099(21)00810-0 PubMed PMID: 35240041.

10. QIAGEN. QuantiFERON®-TB Gold Plus (QFT®-Plus) ELISA Package Insert: The whole blood IFN-γ test measuring responses to ESAT-6 and CFP-10 peptide antigens [Internet]. QIAGEN, Hilden, Germany. Available from: www.QuantiFERON.com

11. Petruccioli E, Chiacchio T, Pepponi I, Vanini V, Urso R, Cuzzi G, et al. First characterization of the CD4 and CD8 T-cell responses to QuantiFERON-TB Plus. J Infect. 2016;73:588–97. doi:10.1016/j.jinf.2016.09.008

12. Ledesma JR, Ma J, Zheng P, Ross JM, Vos T, Kyu HH. Interferon-gamma release assay levels and risk of progression to active tuberculosis: a systematic review and dose-response meta-regression analysis. BMC Infect Dis. 2021;21:467. doi:10.1186/s12879-021-06141-4

13. Barcellini L, Borroni E, Brown J, Brunetti E, Campisi D, Castellotti PF, et al. First evaluation of QuantiFERON-TB Gold Plus performance in contact screening. Eur Respir J. 2016;48:1411–9. doi:10.1183/13993003.00510-2016 PubMed PMID: 27390280.

14. Darmawan G, Liman LMS, Hamijoyo L, Atik N, Alisjahbana B, Sahiratmadja E. Comparison of interferon-gamma production between TB1 and TB2 tubes of QuantiFERON-TB Gold Plus: a meta-analysis. Clin Chem Lab Med CCLM. 2023;61:2067–75. doi:10.1515/cclm-2023-0293

15. Sunshine J, Shaffer M, Han LL, Gaikwad D, Houana AA, Gler MT, et al. Associations Between QuantiFERON-TB Gold Plus IFNγ Concentrations and Progression to Symptomatic Tuberculosis in Global High-Burden TB Settings [Internet]. bioRxiv; 2026 [cited 2026 Feb 7]. p. 2026.01.29.702660. Available from: https://www.biorxiv.org/content/10.64898/2026.01.29.702660v1 doi:10.64898/2026.01.29.702660

16. Metcalfe JZ, Cattamanchi A, McCulloch CE, Lew JD, Ha NP, Graviss EA. Test Variability of the QuantiFERON-TB Gold In-Tube Assay in Clinical Practice. Am J Respir Crit Care Med. 2013;187:206–11. doi:10.1164/rccm.201203-0430OC

17. Amofa-Sekyi M, Schaap A, Mureithi L, Kosloff B, Cheeba M, Kangololo B, et al. Comparing patterns of recent and remote Mycobacterium tuberculosis infection determined using the QuantiFERON-TB Gold Plus assay in a high TB burden setting. PLOS Glob Public Health. 2024;4:e0003182. doi:10.1371/journal.pgph.0003182

18. Phiri MD, Rickman HM, Mbale H, Feasey HRA, Nliwasa M, Schwalb A, et al. Drivers of sex differences in Mycobacterium tuberculosis immunoreactivity among adolescents and adults in Blantyre, Malawi [Internet]. Research Square; 2026 [cited 2026 Jan 17]. Available from: https://www.researchsquare.com/article/rs-8474694/v1 doi:10.21203/rs.3.rs-8474694/v1

19. Rickman HM, Phiri MD, Feasey HRA, Krutikov M, Shao H, Horton KC, et al. Sex differences in the risk of *Mycobacterium tuberculosis* infection: a systematic review and meta-analysis of population-based immunoreactivity surveys. Lancet Public Health. 2025;10:e588–98. doi:10.1016/S2468-2667(25)00120-3

20. Horton KC, MacPherson P, Houben RMGJ, White RG, Corbett EL. Sex Differences in Tuberculosis Burden and Notifications in Low- and Middle-Income Countries: A Systematic Review and Meta-analysis. PLOS Med. 2016;13:e1002119. doi:10.1371/journal.pmed.1002119

21. Feasey HRA, Khundi M, Soko RN, Nightingale E, Burke RM, Henrion MYR, et al. Prevalence of bacteriologically-confirmed pulmonary tuberculosis in urban Blantyre, Malawi 2019–20: Substantial decline compared to 2013–14 national survey. PLOS Glob Public Health. 2023;3:e0001911. doi:10.1371/journal.pgph.0001911

22. Rickman HM, Phiri MD, Feasey HRA, Mbale H, Nliwasa M, Semphere R, et al. Tuberculosis Immunoreactivity Surveillance in Malawi (Timasamala)—A protocol for a cross-sectional Mycobacterium tuberculosis immunoreactivity survey in Blantyre, Malawi. PLOS ONE. 2024;19:e0291215. doi:10.1371/journal.pone.0291215

23. Middelkoop K, Bekker LG, Liang H, Aquino LD, Sebastian E, Myer L, et al. Force of tuberculosis infection among adolescents in a high HIV and TB prevalence community: a cross-sectional observation study. BMC Infect Dis. 2011;11:156. doi:10.1186/1471-2334-11-156

24. R Core Team. R: A Language and Environment for Statistical Computing [Internet]. Vienna, Austria: R Foundation for Statistical Computing; 2024 [cited 2024 Nov 23]. Available from: https://www.R-project.org/

25. Bürkner PC. brms: An R Package for Bayesian Multilevel Models Using Stan. J Stat Softw. 2017;80:1–28. doi:10.18637/jss.v080.i01

26. Vehtari A, Gelman A, Gabry J. Practical Bayesian model evaluation using leave-one-out cross-validation and WAIC. Stat Comput. 2017;27:1413–32. doi:10.1007/s11222-016-9696-4

27. Ministry of Health. National Tuberculosis Control Programme Manual: Eight Edition. Ministry of Health, Lilongwe, Malawi; 2017.

28. Black GF, Weir RE, Chaguluka SD, Warndorff D, Crampin AC, Mwaungulu L, et al. Gamma Interferon Responses Induced by a Panel of Recombinant and Purified Mycobacterial Antigens in Healthy, Non-Mycobacterium bovis BCG-Vaccinated Malawian Young Adults. Clin Vaccine Immunol. 2003;10:602–11. doi:10.1128/CDLI.10.4.602-611.2003

29. Crampin AC, Glynn JR, Fine PEM. What has “Karonga” taught us? Tuberculosis studied over three decades. Int J Tuberc Lung Dis Off J Int Union Tuberc Lung Dis. 2009;13:153–64. PubMed PMID: 19146741; PubMed Central PMCID: PMC3272402.

30. MacPherson P, Khundi M, Nliwasa M, Choko AT, Phiri VK, Webb EL, et al. Disparities in access to diagnosis and care in Blantyre, Malawi, identified through enhanced tuberculosis surveillance and spatial analysis. BMC Med. 2019;17:21. doi:10.1186/s12916-019-1260-6

31. Whittaker E, Nicol MP, Zar HJ, Tena-Coki NG, Kampmann B. Age-related waning of immune responses to BCG in healthy children supports the need for a booster dose of BCG in TB endemic countries. Sci Rep. 2018;8:15309. doi:10.1038/s41598-018-33499-4

32. Lewinsohn DA, Lewinsohn DM, Scriba TJ. Polyfunctional CD4+ T Cells As Targets for Tuberculosis Vaccination. Front Immunol. 2017;8. doi:10.3389/fimmu.2017.01262

33. Bini EI, Mata Espinosa D, Marquina Castillo B, Barrios Payán J, Colucci D, Cruz AF, et al. The Influence of Sex Steroid Hormones in the Immunopathology of Experimental Pulmonary Tuberculosis. PLoS ONE. 2014;9:e93831. doi:10.1371/journal.pone.0093831 PubMed PMID: 24722144; PubMed Central PMCID: PMC3983091.

34. Viana Machado F, Morais C, Santos S, Reis R. Evaluation of CD8+ response in QuantiFERON-TB Gold Plus as a marker of recent infection. Respir Med. 2021;185:106508. doi:10.1016/j.rmed.2021.106508

35. Chee CBE, Kyi-Win K, Tan S, Wang YT. QuantiFERON-TB Gold Plus CD8+ T cell responses in contacts with tuberculosis disease and recent tuberculosis infection. Obar JJ, editor. Microbiol Spectr. 2025;13:e01353–25. doi:10.1128/spectrum.01353-25

36. Pérez-Recio S, Pallarès N, Grijota-Camino MD, Sánchez-Montalvá A, Barcia L, Campos-Gutiérrez S, et al. Identification of Recent Tuberculosis Exposure Using QuantiFERON-TB Gold Plus, a Multicenter Study. Microbiol Spectr. 2021;9:e00972–21. doi:10.1128/Spectrum.00972-21

37. Oh CE, Ortiz-Brizuela E, Bastos ML, Menzies D. Comparing the Diagnostic Performance of QuantiFERON-TB Gold Plus to Other Tests of Latent Tuberculosis Infection: A Systematic Review and Meta-analysis. Clin Infect Dis. 2021;73:e1116–25. doi:10.1093/cid/ciaa1822

