## Supplemental File for "Quantitative interferon gamma responses to *Mycobacterium tuberculosis* in a community-based survey of adolescents and adults in Blantyre, Malawi"

### Supplementary material

**Supplementary Figure 1: QuantiFERON-TB Gold Plus nil-subtracted TB1 and TB values by age and sex**

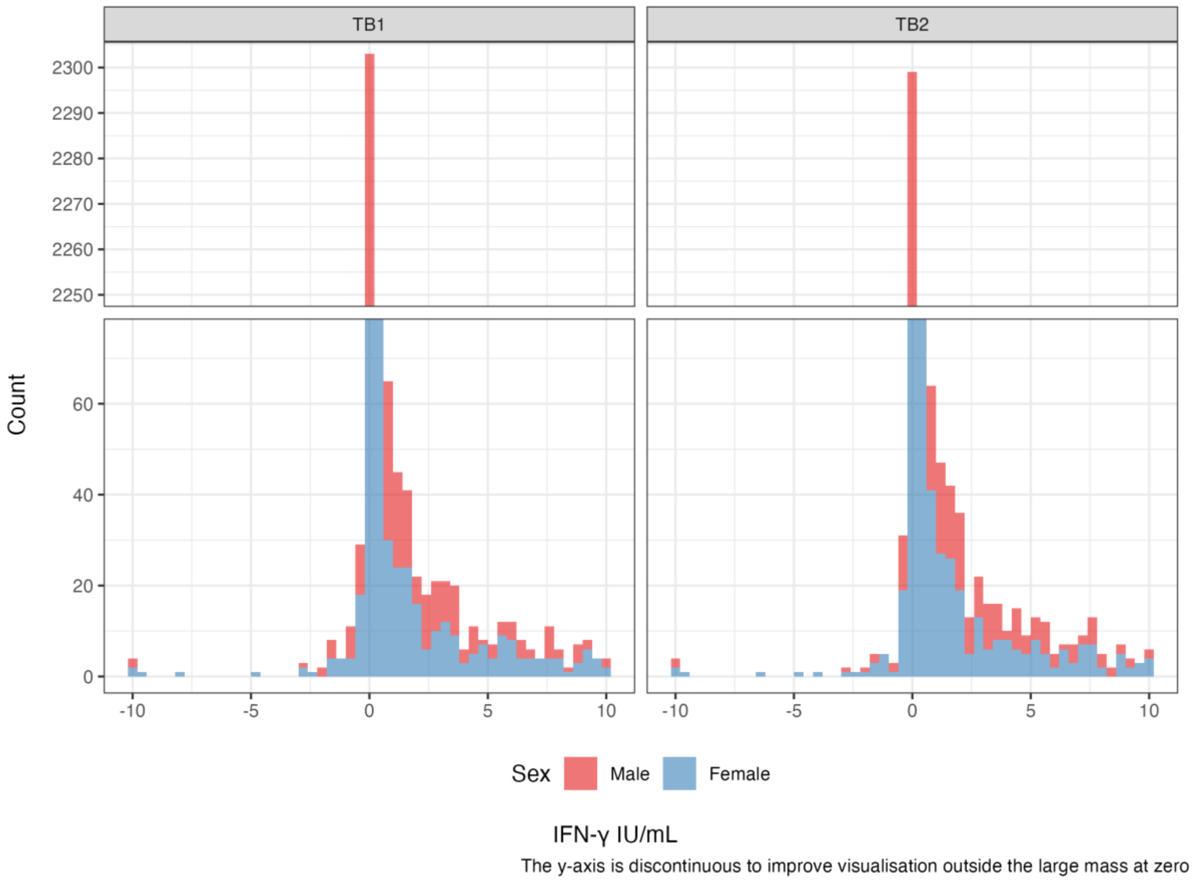


Histogram showing the distributions of QuantiFERON-TB Gold Plus nil-subtracted TB1 and TB2 values by age and sex. IFN-γ: interferon gamma. IU/mL: international units per millilitre. Note: IFN-γ values are right-truncated at the assay upper limit of detection (10 IU/mL).

**Supplementary Table 1: Summary values for QuantiFERON-TB Gold Plus nil-subtracted TB1 and TB2 values**

| Category (IU/mL) | TB1 % (n/N) | TB2-Nil (n/N) |
|---|---|---|
| **<0.00** | 25.3 (731/2895) | 23.8 (689/2895) |
| **0.00** | 22.1 (641/2895) | 19.2 (555/2895) |
| **<0.35** | 84.6 (2448/2895) | 84.1 (2435/2895) |
| **≥0.35** | 15.4 (447/2895) | 15.9 (460/2895) |
| **<0.2** | 81.5 (2360/2895) | 8 1.2 (2350/2895) |
| **0.2-0.7** | 6.4 (186/2895) | 6.1 (178/2895) |
| **>0.7-10** | 12.1 (349/2895) | 12.7 (367/2895) |
| **>10.00[1]** | 2.1 (60/2895) | 2.0 (57/2895) |

1. Original values reported by QuantiFERON-TB Gold Plus software as “>10 IU/mL” before nil subtraction. These values were recoded to “10 IU/mL” for inclusion in quantitative analyses.

**Supplementary Table 2: Correlation of responses estimated from the joint multivariate normal distribution of nil-subtracted QuantiFERON Gold-TB Gold Plus TB1 and TB2 responses**

| Characteristic | Correlation parameter[1], ρ (95% CrI) |
|---|---|
| **Overall** | 0.86 (0.85-0.87) |
| **Age groups (years)** | |
| 10-19 | 0.83 (0.81-0.84) |
| 20-29 | 0.86 (0.85-0.88) |
| 30-40 | 0.88 (0.87-0.90) |
| **Sex** | |
| Male | 0.82 (0.80-0.84) |
| Female | 0.89 (0.88-0.90) |

1. Correlation estimates are from unadjusted multivariate normal models fitted to TB1 and TB2, fitted to all observations (overall) or separately for each characteristic value.

**Supplementary Figure 2: Posterior predictive distributions for QuantiFERON Gold-TB Gold Plus nil-subtracted TB1 and TB2 responses**

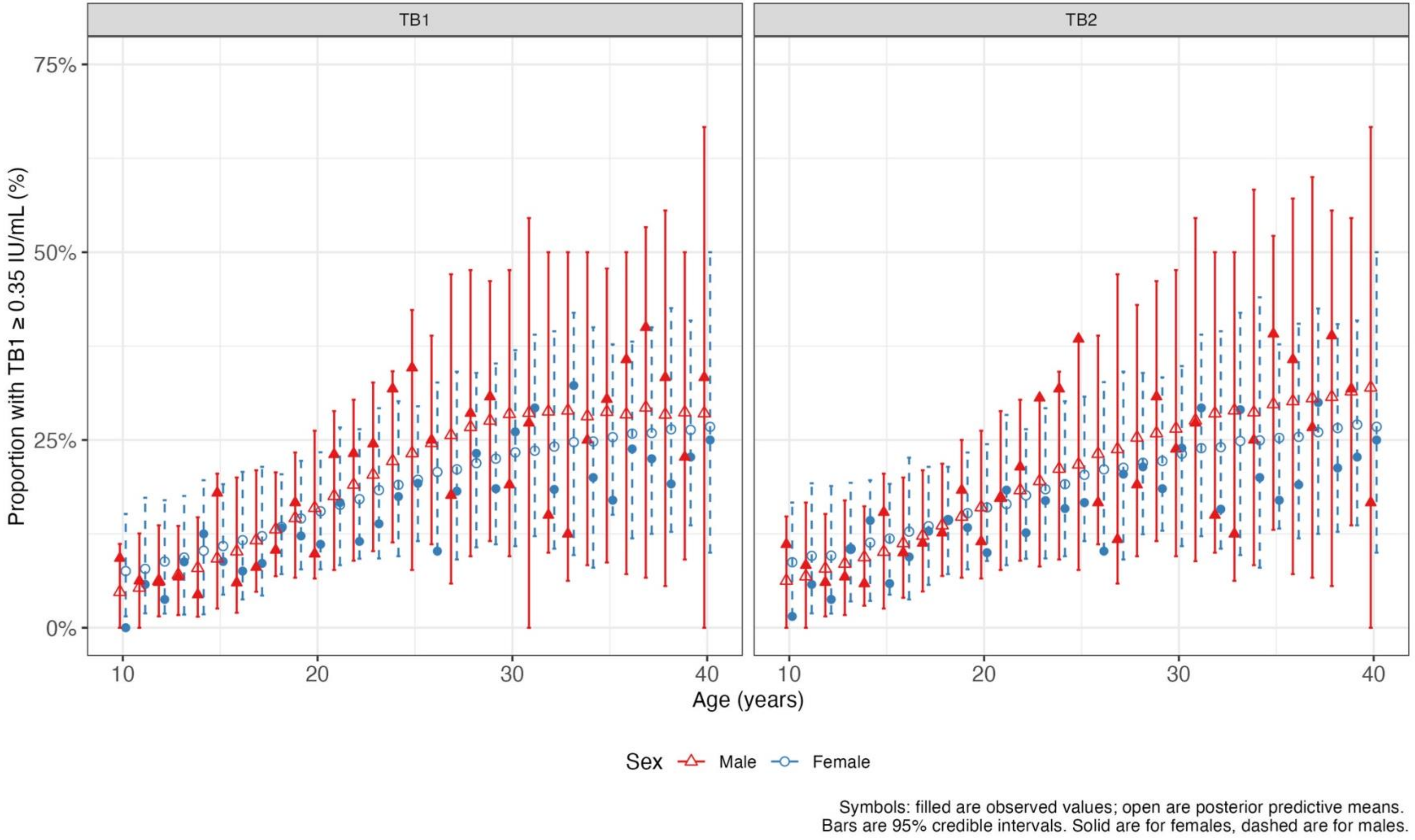

**Supplementary Figure 3: Distribution of QuantiFERON Gold-TB Gold Plus nil-subtracted TB2 minus TB1 differential**

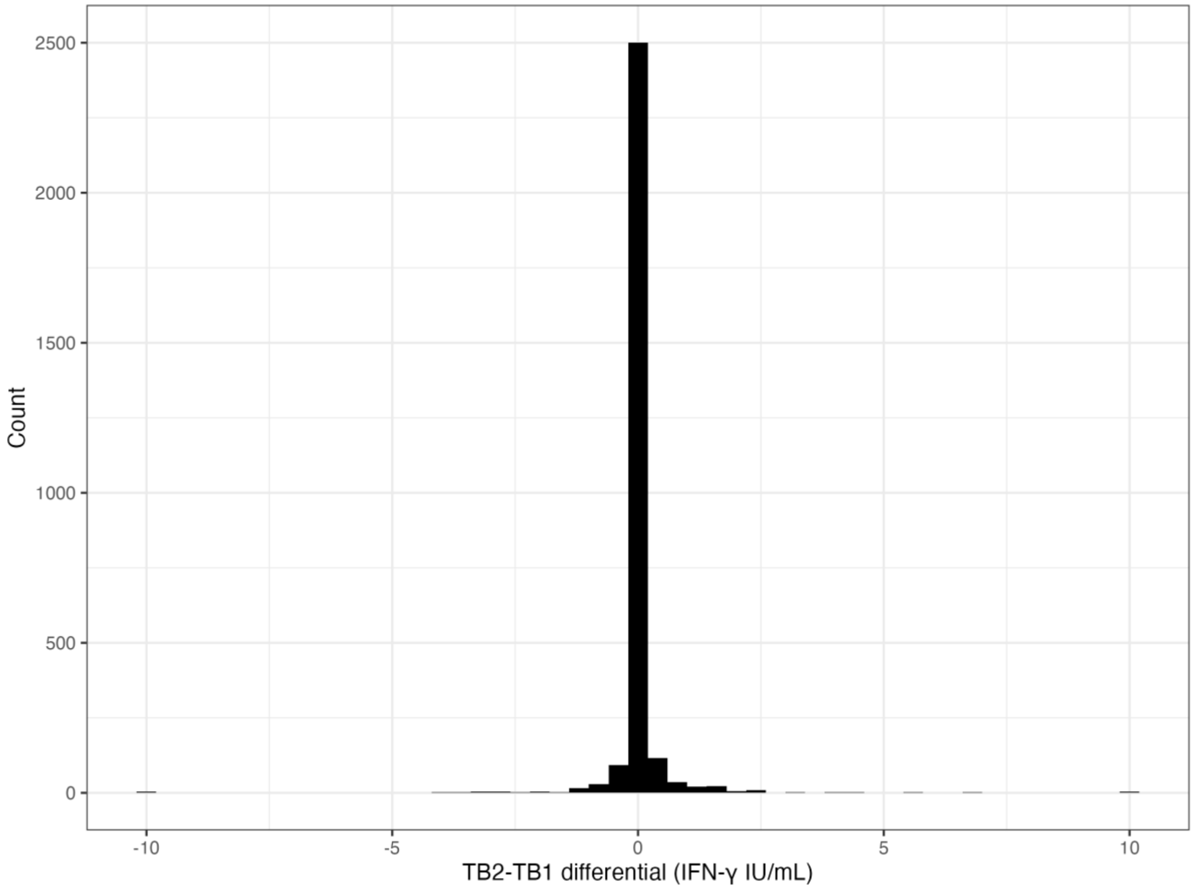


Histogram showing the distributions of QuantiFERON-TB Gold Plus nil-subtracted TB2 minus TB1 differential. IFN-γ: interferon gamma. IU/mL: international units per millilitre.

**Supplementary Table 3: Factors associated with the magnitude of nil-subtracted QuantiFERON Gold-TB Gold Plus TB1 and TB2 responses[1] at a theoretical 0.1 IU/mL positivity cut-off**

| Covariate | TB1 | | TB2 | |
|---|---|---|---|---|
| | Univariable effect[3] (95% CrI) | Multivariable effect[2] (95% CrI) | Univariable effect[3] (95% CrI) | Multivariable effect[2] (95% CrI) |
| HIV positive | 0.88 (0.56-1.37) | 0.86 (0.55-1.37) | 1.22 (0.73-2.05) | 1.19 (0.71-2.09) |
| Previous TB[4] treatment | 1.25 (0.68-2.24) | 1.24 (0.61-2.56) | 1.22 (0.68-2.19) | 1.28 (0.64-2.47) |
| Previous household TB contact[5] | 1.01 (0.70-1.45) | 1.08 (0.69-1.66) | 1.08 (0.75-1.54) | 1.06 (0.69-1.60) |
| Mean intercept (exponentiated scale)[6] | - | 2.45 (2.04-3.06) | - | 2.47 (2.06-3.09) |
| Sigma (log scale)[6] | - | 1.16 (1.03-1.31) | - | 1.16 (1.03-1.31) |
| Hurdle intercept[7] (probability scale) | 0.84 (0.83-0.86) | 0.81 (0.79-0.83) | 0.84 (0.82-0.85) | 0.80 (0.78-0.82) |

1. Bayesian hurdle lognormal models were fitted separately to TB1 and TB2 responses. Values <0.10 IU/mL were treated as “non-response", and recoded to 0 IU/mL for model fitting purposes.
2. Univariable effect (exponentiated scale) and 95% credible interval (CrI) on the mean log response estimated from lognormal components of hurdle lognormal models.
3. Multivariable effect and 95% credible interval (exponentiated scale) on the mean log response estimated from lognormal components of hurdle lognormal models adjusted for all covariates included in the table.
4. TB: Tuberculosis.
5. Household TB contact was defined as living in the same household with a person treated for TB.
6. Intercept and sigma (both log scale) parameters of the lognormal components from multivariable models.
7. Intercept parameter of the hurdle component, estimating the proportion of responses <0.35 IU/mL.
